# Metacognition in Functional Cognitive Disorder

**DOI:** 10.1101/2021.06.24.21259245

**Authors:** Rohan Bhome, Andrew McWilliams, Gary Price, Norman A Poole, Robert J Howard, Stephen M Fleming, Jonathan D Huntley

## Abstract

Functional cognitive disorder (FCD) is common but underlying mechanisms remain poorly understood. Metacognition, an individual’s ability to reflect on and monitor cognitive processes, is likely to be relevant. Local metacognition refers to an ability to estimate confidence in cognitive performance on a moment-to-moment basis, whereas global metacognition refers to long-run self-evaluations of overall performance. Using a novel protocol comprising task-based measures and hierarchical Bayesian modelling, we compared local and global metacognitive performance in individuals with FCD and evaluated interactions between these levels of metacognition. We also investigated how local and global metacognition were related to the presence of affective symptomatology.

Eighteen participants with FCD were recruited to this cross-sectional study. Participants completed computerised tasks that enabled local metacognitive efficiency for perception and memory to be measured using the hierarchical meta-d’ (HMeta-d) model within a signal detection theory framework. Participants also completed the Multifactorial Memory Questionnaire (MMQ) measuring global metacognition (beliefs about memory performance), and questionnaires measuring anxiety and depression. Estimates of local metacognitive efficiency were compared to those estimated from two control groups who had undergone comparable metacognitive tasks. Global metacognition scores were compared to existing normative data. A hierarchical regression model was used to evaluate associations between global metacognition, depression and anxiety and local metacognitive efficiency, while simple linear regressions were used to evaluate whether affective symptomatology and local metacognitive confidence were associated with global metacognition.

Participants with FCD had intact local metacognition for perception and memory when compared to controls, with the 95% highest-density intervals for metacognitive efficiency overlapping with the two control groups in both cognitive domains. FCD participants had significantly lower global metacognition scores compared to normative data; MMQ-Ability (t=6.54, p<0.0001) and MMQ-Satisfaction (t=5.04, p<0.0001). Mood scores, global metacognitive measures and metacognitive bias were not significantly associated with local metacognitive efficiency. Increased local metacognitive bias (β= −0.20 (SE=0.09), q= 0.01) and higher depression scores as measured by Patient Health Questionnaire-9 (β= −1.40 (SE=2.56), q= 0.01) were associated with lower global metacognition scores.

We show that local metacognition is intact, whilst global metacognition is impaired, in FCD, suggesting a decoupling between the two metacognitive processes. In a Bayesian model, an aberrant prior (impaired global metacognition), may override bottom up sensory input (intact local metacognition), giving rise to the subjective experience of abnormal cognitive processing. Future work should further investigate the interplay between local and global metacognition in FCD and aim to identify a therapeutic target to recouple these processes.

## Introduction

Functional Cognitive Disorder (FCD) is an increasingly recognised condition, and is seen in up to 25% of patients in cognitive assessment services.^1^ FCD is characterised by persistent and distressing cognitive symptoms that cause significant difficulty and cannot be explained by another disorder.^1,2^ A canonical diagnostic feature is cognitive internal inconsistency, which refers to a discrepancy between the individual’s subjective appraisal of their cognitive performance compared to objective evidence about that performance, as well as differences in cognitive performance under automatic and explicit control.^2^ Metacognition, the ability of an individual to reflect on and monitor their own cognitive processes,^3,4^ has been posited as a key mechanism contributing to cognitive internal inconsistency.^1,5-8^

The exact role of metacognitive processing in FCD remains poorly understood. Theoretical frameworks distinguish between two aspects of metacognition, global and local, and evidence from behavioural experiments suggests that performance in one can modulate the other.^9^ Global metacognition refers to self-performance estimates (SPEs) about overall task performance, and in FCD these may be shaped by self-evaluation of cognitive performance, emotions associated with cognitive ability and perceived control over cognitive functioning.^7,10^ Global metacognition, measured using subjective rating tools, has been shown to be altered in patients with FCD.^7,11,12^

Local metacognition - the ability to track changes in moment-to-moment cognitive performance - is focused on the appraisal of individual cognitive decisions as opposed to evaluation of overall cognitive performance. Local metacognition can be characterised by two key parameters – metacognitive bias and metacognitive sensitivity. Metacognitive bias describes an individual’s average confidence level in their task performance, regardless of whether their judgments are objectively correct or incorrect – and therefore overlaps conceptually with notions of global metacognition. In contrast, metacognitive sensitivity refers to the ability to distinguish between correct and incorrect decisions using confidence ratings. This construct can be objectively quantified through a combination of task-based measures and type 2 signal detection theory.^13-15^ However, metrics of metacognitive sensitivity are known to be confounded by variation in first-order cognitive performance and by an individual’s bias to give generally positive or negative confidence ratings.^13^ This has led to the development of model-based measures of metacognitive “efficiency”, such as meta-d’/d’, which characterise the level of metacognitive sensitivity relative to a particular level of task performance.^15^ Local metacognitive efficiency is likely to be relevant in FCD for two reasons. First, it is considered to play a role in directly modulating global metacognition which,^9^ as mentioned above, is itself distorted in FCD.^7,11-12^ Second, it likely regulates the expression of core symptoms of FCD, including increased but poorer quality monitoring for cognitive lapses,^6-7^ and misinterpretation of what may in fact be attentional lapses.^16^

In this study, we evaluated and compared global and local metacognition in FCD participants. Whilst global metacognition has previously been evaluated in people with FCD,^7,11-12^ ours is the first study to objectively measure local metacognition. We did this using a novel protocol comprising task-based measures and hierarchical Bayesian modelling.^13,17-18^ We compared local metacognition in FCD with two healthy control groups constructed from previously collected data.^19,20^ In addition, we investigated whether depression and anxiety were associated with measures of global and local metacognition. Our primary hypothesis was that global and local metacognition are coupled and so both would be impaired in people with FCD. Secondarily, we hypothesised that depression and anxiety may predict lower global self-confidence when giving metacognitive ratings, because the negative cognitions that arise in these conditions^21^ would affect individuals’ perception of their cognitive performance.

## Materials and Methods

### Participants

Eighteen participants (ages 24-66 years, mean 49.22 (SD= 13.55); 8 females) with an established diagnosis of FCD were recruited from two tertiary neuropsychiatric clinical services (National Hospital for Neurology and Neurosurgery (NHNN) (University College London Hospitals NHS Trust) and South West London and St. George’s Mental Health Trust). Participants provided informed consent which was obtained according to the Declaration of Helsinki principles for medical research.

### Inclusion Criteria

Inclusion criteria were age above 18, a diagnosis of FCD and provision of informed consent to participate in the study. The diagnosis of FCD was made by a tertiary specialist multidisciplinary team based on the following criteria;^2^ a) the presence of one or more symptoms of impaired cognitive function causing clinically significant distress and/or impairment in functioning, b) evidence of cognitive internal inconsistency and c) symptoms of impaired cognitive functioning that cannot be accounted for by an alternative medical condition. Patients with a diagnosis of a neurodegenerative disorder on the basis of neuroimaging, neuropsychological and clinical assessment were excluded.

### Comparison groups

One group (Control Group 1)^19^ was drawn retrospectively from a large (n=304) web study of healthy volunteers, recruited via the academic crowdsourcing site, Prolific (https://www.prolific.co/). Controls were matched by age alone, irrespective of gender, to the patient group, in the ratio of 3:1 respectively. The same tasks were undertaken as those completed by the FCD group, with an identical staircasing procedure used to control for first-order task performance, although there were some minor differences between task versions for the two groups, as the FCD group completed an earlier version of the tasks (see below).

A second control group (Control Group 2)^20^ was formed by re-analysing previously published behavioural data from a neuroimaging study of healthy volunteers at New York University, undertaking tasks of a similar psychophysical structure to our own.

### Local metacognition

A task measuring metacognition for memory and a task measuring metacognition for perception (vision) were administered. Each contained 5 mini-blocks of 20 trials, with each trial involving a two-alternative forced-choice (2-AFC) task followed immediately by a metacognitive (confidence) rating, in line with standard task structures used in previous research.^20^ An optional set of five practice trials before each set of 20 trials allowed participants to familiarise themselves with the first-order task and the confidence rating sliding scale.

In each recognition memory trial, an array of stimuli is presented for two seconds for memorisation. A novel stimulus is then presented together with a stimulus from the memorisation set, and the participant is asked to indicate which stimulus they had memorised. The subject is then asked, “How confident are you?” and must make a metacognitive judgement using a sliding scale, which is marked with the labels at either end of “complete guess” and “absolutely certain” (**Fig. 1**). Task difficulty is staircased to increase subject motivation and control first-order performance, by increasing or decreasing the number of items in the stimulus sets presented for memorisation. The staircase aims to keep task accuracy for each subject around 70-72%, by adjusting difficulty in response to changes in first-order task performance in line with established protocols.^22^ When a subject gives an incorrect response, the task is made easier by one increment (decreasing by one the size of the memory stimulus set presented for memorisation). When a subject gives two consecutive correct responses, the task is made more difficult by one increment (increasing by one the size of the memory stimulus set). The memory task undertaken by the FCD group employed stimuli which were less visually appealing, with plain white, rather than coloured, stimuli. For each of the 5 memory task themes, there were 12 available stimuli for display to the FCD group, whereas the version used by the control group had 25 stimuli within each task theme.

**Figure 1.**
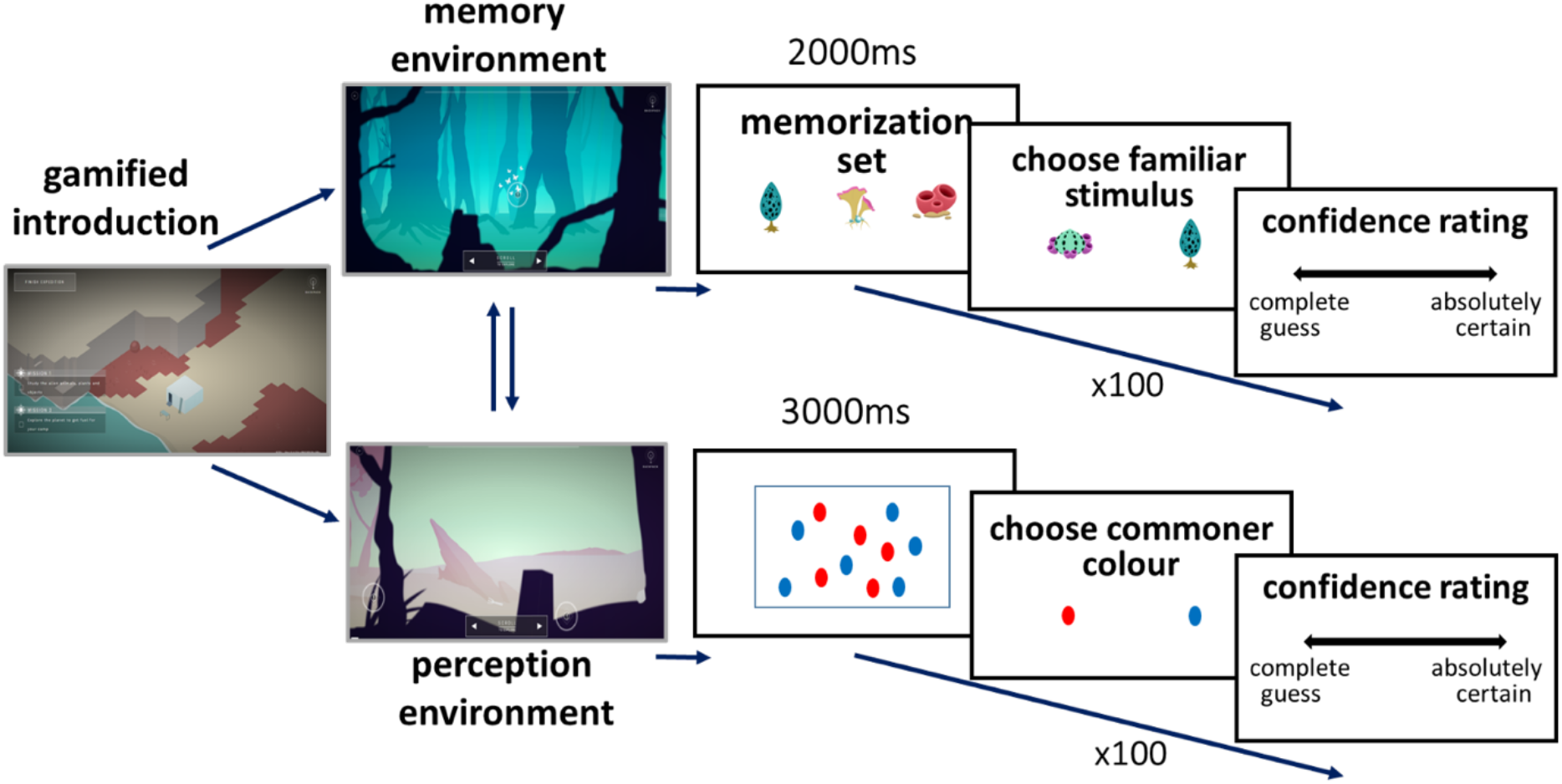
Task structure. Participants were tested on both recognition memory (left) and perceptual discrimination (right) tasks. In both tasks, participants are: 1) presented stimuli (for 2000ms in the recognition memory task; 3000ms in the perceptual discrimination task; 2) asked to make an unspeeded two-alternative forced-choice based on presented stimuli and 3) asked to make an unspeeded metacognitive judgement on a sliding scale.

In each perceptual discrimination trial, the subject is presented with an array of identical red and blue shapes for three seconds. Subjects are then asked whether there were more red or blue shapes. They then undertake the same metacognitive rating judgement as in the memory task, before moving onto the next perceptual trial (**Fig. 1**). The version given to the FCD group used less visually appealing stimuli and a first trial presenting 41 shapes of one colour and 25 of the other, whereas the control group were shown 40 and 25. Difficulty was staircased by increasing or decreasing this difference, again aiming to converge the first-order task accuracy of each subject to 70-72%.^22^ When a subject gives an incorrect response, the task is made easier by one increment (increasing the difference by one, through increasing the larger number of coloured shapes presented). When a subject gives two consecutive correct responses, the task is made more difficult by one increment (decreasing the difference by one, through decreasing the larger number of coloured shapes presented).

The first 20 trials from each task (memory and perception) were removed prior to analysis to allow for learning effects and the difficulty staircase used to calibrate task difficulty to stabilise. Trials where subjects took longer than 30 seconds to respond to any instruction were also removed.

### Self-reported measures

Global metacognition was measured using the Multifactorial Memory Questionnaire (MMQ).^23^ The MMQ has three subscales; MMQ-Ability measuring self-appraisal of memory ability, MMQ-Satisfaction measuring satisfaction and concern about memory and MMQ-Strategy which measures the use of compensatory strategies and cognitive aids in daily functioning. Lower scores in the first two subscales suggest lower global memory self-performance estimates (SPEs) whilst higher scores in the MMQ-Strategy subscale indicate greater subjective use of memory strategies.

The Patient Health Questionnaire-9 (PHQ-9)^24^ and the Generalised Anxiety Disorder-7 (GAD-7)^25^ scale were used to measure symptoms of depression and anxiety respectively.

### Statistical analysis

IBM SPSS Version 22 was used to calculate descriptive statistics for study variables including age, MMQ subscale scores, PHQ-9 and GAD-7. The MMQ subscale scores in our study for FCD patients were compared to normative data^26^ using independent t-tests.

To analyse local metacognitive data, the meta-d’ model was used to calculate metacognitive efficiency within a signal detection theoretic framework.^15^ Meta-d’ reflects an individual’s metacognitive sensitivity, namely how well a subject discriminates correct from incorrect responses. The meta-d’/d’ ratio (M-ratio) is known as metacognitive efficiency, and quantifies metacognitive sensitivity (meta-d’) relative to task performance (d’). An optimal value for metacognitive efficiency is therefore 1. Local metacognitive efficiency was calculated using a hierarchical Bayesian model as implemented within the HMeta-d toolbox^18^ in JAGS. This toolbox was developed with the aim of allowing robust estimates of local metacognitive efficiency in situations where limited per-participant task data are available, such as in clinical populations. Hierarchical Bayesian modelling was used to generate group-level parameter estimates, thereby allowing direct inference on group comparisons (such as patients vs. controls) while avoiding reliance on noisy point estimates of single-subject parameters.^18^ Certainty on these parameters (the group-level M-ratios), was determined by computing the 95% high-density interval (HDI) from the posterior samples.^27^ Metacognitive efficiency at the group level could thus be compared with existing data from healthy controls by computing the HDIs of the differences between the posterior estimates obtained from each dataset.

An extended version of the HMeta-d model (the RHMeta-d model) has been developed to hierarchically estimate regression parameters relating metacognitive efficiency to covariates of interest.^28^ Whilst capitalising on the power of hierarchical estimation, the extended model avoids the problems encountered by post-hoc regressions on hierarchical model parameters such as unwanted shrinkage to the group mean. This model was used to regress MMQ, PHQ-9 and GAD-7 scores on memory M-ratio. Although the hierarchical regression model was able to return a robust characterisation of influences on metacognitive efficiency at the group level, the small numbers of trials (N=80) completed by each individual subject meant that it was not possible to reliably identify individual-level M-ratios.

Associations of demographic characteristics, affective symptomatology and other metacognitive measures with global metacognition, measured using MMQ-Ability, were evaluated using simple linear regression models. Covariates were not included because age and gender are known to not significantly affect MMQ subscale scores.^26^ While measured and interpreted individually, for theoretical reasons there is likely to be a strong correlation between different MMQ subscales and so these were not included as covariates. A false discovery rate (FDR) correction was performed for the six dependent variables included in these analyses, using the Benjamini-Hochberg method.^29^

The M-ratio for memory was chosen over the M-ratio for perception in these analyses because the MMQ is most focussed on symptoms of memory impairment - rather than symptoms concerning other types of cognition - thereby enabling a domain-specific evaluation of the interplay between local and global metacognition.

### Missing Data

For particular metacognitive tasks or questionnaire measures, participants were excluded if they could not tolerate or complete testing and this is highlighted in the results section. Analyses were performed on available data without imputation.

### Ethical Approval

Ethical approval (ref: 18/LO/1056) from NHS Health Research Authority (HRA) and Health and Care Research Wales (HCRW) and London-South East Research Ethics Committee (REC) was granted to carry out this study using FCD patients. Ethical approval for collection of data from healthy volunteers was given by the University College London Ethics Committee (ref: 12/0006).

### Data Availability

Data used in this study will be shared upon reasonable request to the corresponding author. All data and statistics generated from this study are presented.

## Results

### Demographics, depression and anxiety scores

Of the eighteen participants in this study, 8 (44%) were female. The average age was 49.2 years (SD 13.6, range 24-62) (**Table 1**). Based on validated PHQ-9 cut-offs, 14 (82%) were in the depressed range, three (18%) with mild symptoms, seven (41%) moderate, three (18%) moderately severe and one (6%) severe. Based on GAD-7 cut-offs, 10 (59%) of the participants were in the anxious range-seven (41%) with mild symptoms, two (12%) moderate and one (6%) severe symptoms of anxiety. Control Group 1^19^ was matched for age and consisted of 54 individuals, with mean age of 49.1 years (SD 13.3, range 24-62). Control Group 2^20^ participants were significantly younger (n=30, mean 24.97, SD=4.44, range 18-33) in comparison to the FCD study population (t=9.04, df=46, p<0.001).

**Table 1.** Demographics, mood and global metacognitive characteristics of FCD participants compared to normative sample

Demographics, depression and anxiety scores
| Attribute | FCD participants (n=18) |  | Normative data<sup>a</sup> (n=401) |  | Statistic<sup>b</sup> |
| --- | --- | --- | --- | --- | --- |
|  | Mean (SD) | Range | Mean (SD) | Range |  |
| Age | 49.2 (13.6) | 24-66 | 71.4 (8.9) | 39-91 | t=10.08, df=417, p<0.0001 |
| Gender, male, n (%) | 10 (56) | | 116 (29) | | $\chi^2=5.81$, p=0.02 |
| PHQ-9<sup>c</sup> | 11.5 (5.6) | 3-25 |  |  |  |
| GAD-7<sup>c</sup> | 7.0 (4.8) | 0-20 |  |  |  |
| MMQ- Satisfaction<sup>c</sup> | 26.9 (11.9) | 12-56 | 43.9 (13.7) | 7-72 | t=5.04, df=416, p<0.0001 |
| MMQ- Ability<sup>c</sup> | 30.4 (14.7) | 12-50 | 48.8 (11.2) | 0-80 | t=6.54, df=416, p<0.0001 |
| MMQ- Strategy<sup>c</sup> | 36.8 (11.9) | 19-57 | 37.3 (10.4) | 1-64 | t=0.19, df= 416, p=0.85 |
| Abbreviations: GAD-7= Generalised Anxiety Disorder-7; PHQ-9= Patient Health Questionnaire-9; MMQ= Multifactorial Memory Questionnaire |  |  |  |  |  |
| <sup>a</sup>Normative MMQ data (Troyer and Rich, 2018) |  |  |  |  |  |
| <sup>b</sup>Independent t-tests (MMQ data and age) and $\chi^2$ (gender) were used to compare FCD participants with normative MMQ data | | | | | |
| <sup>c</sup>n=17 |  |  |  |  |  |

### Local Metacognition task

First-order task performance for each group is presented using the signal-detection theory-derived metric d’ (**Table 2**). The minimal inter-subject variability in d’ indicates that task difficult was appropriately titrated for each individual using staircases. Group-level posteriors over M-ratio (meta-d’/d’) were compared between FCD participants and the two control groups (**Fig. 2)**. Analysis of perceptual metacognition data was undertaken for only 14 of the FCD participants: four were excluded for completing too few trials. The means of the posterior densities of group-level M-ratio estimates for FCD participants undertaking both the perceptual (0.62) and memory (1.31) tasks are very close to those found in the healthy Control Group 1 (0.56 and 1.13, respectively) and Control Group 2 (0.66 and 1.36). Highest Density Intervals (HDIs), for the patient and control groups can be seen to be almost coincident, indicating no evidence of a difference in local metacognition between the groups (**Table 2**).

**Table 2.** Measures of local metacognition

Local Metacognition task
| Table 2. Measures of local metacognition |  |  |  |  |  |  |
| --- | --- | --- | --- | --- | --- | --- |
| Perception |  |  |  | Memory |  |  |
| Group | n | d'<sup>a</sup> | M-ratio<sup>b</sup> [HDI] | n | d'<sup>a</sup> | M-ratio<sup>b</sup> [HDI] |
| FCD | 14<sup>c</sup> | 1.26 (0.40) | 0.62 [0.39, 0.87] | 18 | 1.03 (0.19) | 1.31 [1.08, 1.56] |
| Controls 1 | 42 | 1.33 (0.23) | 0.56 [0.43, 0.69] | 54 | 1.14 (0.21) | 1.13 [1.00, 1.26] |
| Controls 2 | 24 | 1.13 (0.35) | 0.66 [0.51, 0.82] | 24 | 1.12 (0.40) | 1.36 [1.17, 1.58] |
| Abbreviations: FCD: Functional Cognitive Disorder; HDI: 95% Highest Density Interval of sampled M-ratios. |  |  |  |  |  |  |
| <sup>a</sup>d', a metric of objective task performance per participant: mean (SD) |  |  |  |  |  |  |
| <sup>b</sup>metacognitive efficiency, calculated as meta- d'/d' : mean (SD) of sampled group parameters |  |  |  |  |  |  |
| <sup>c</sup>n=14 because four participants were unable to complete an adequate number of perception trials |  |  |  |  |  |  |

**Figure 2.**
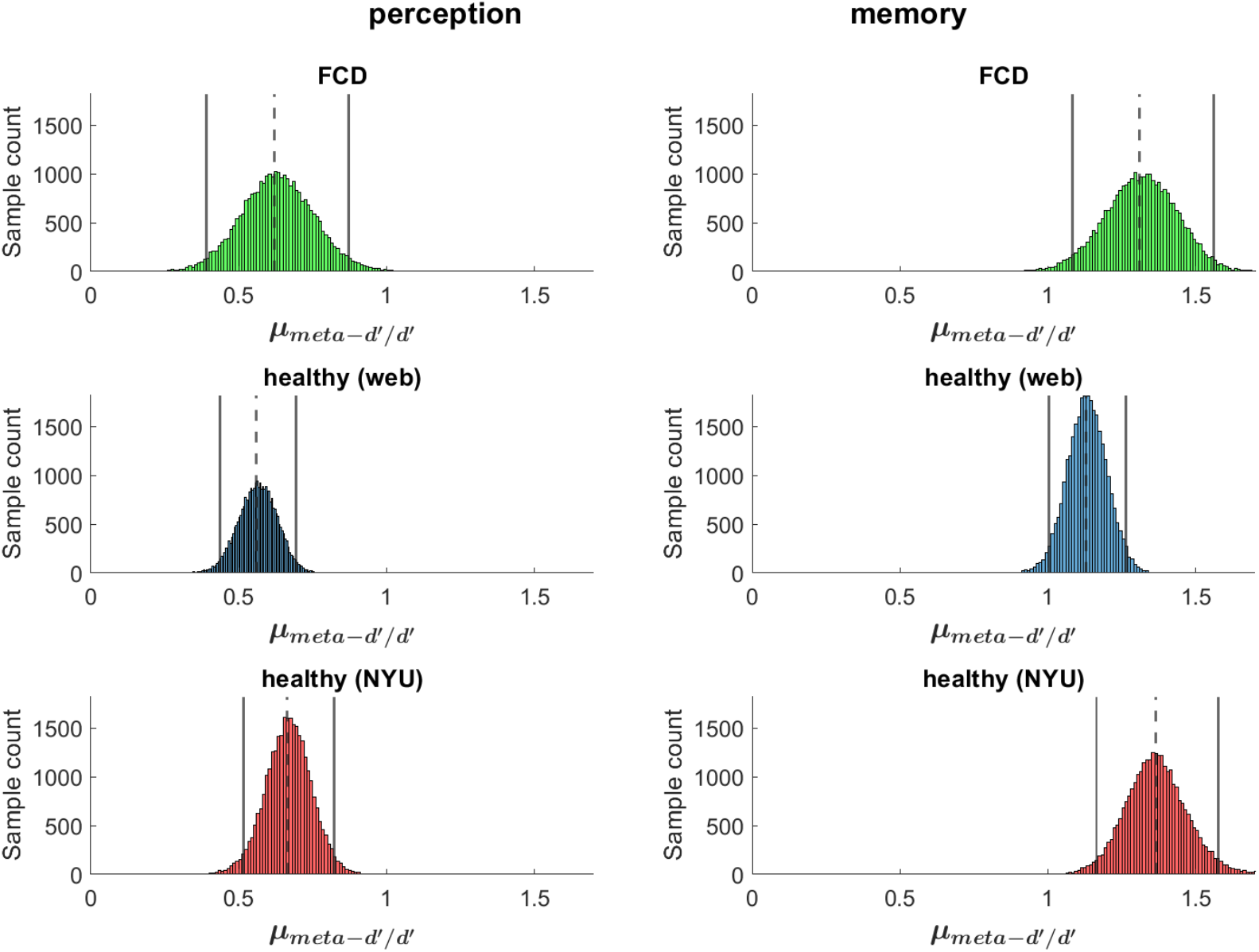
Local metacognition is intact in FCD. Posterior densities of the estimates of group mean metacognitive efficiency (M-ratio) are shown for perception and for memory, in functional cognitive disorder (FCD), Control Group 1 (healthy (web)) and Control Group 2 (healthy (NYU)). Vertical bars show the 95% Highest Density Interval for each M-ratio and dotted vertical bars the mean. Overlap between the HDIs across panels indicates that the FCD group has a similar level of local metacognitive efficiency compared to the controls in both domains.

All three groups had better metacognitive efficiency for memory than for perceptual tasks, consistent with previous findings. Differences in posteriors for the memory and perception tasks were calculated for each group, generating a Bayesian probability that M-ratios differed between the domains and providing strong evidence for a difference in each of the three groups (functional cognitive disorder: *P =* 0.9999; control group 1: *P* = 1; control group 2: *P* = 1).

### Global Metacognition

We compared global metacognition scores in our FCD participants with existing, validated normative data^26^ (**Table 1**). The FCD sample was significantly younger than the normative population (mean= 71.4, SD= 8.9, range= 39-91) (p< 0.001). Mean MMQ-Ability, a surrogate for global SPEs, was significantly lower in our study participants with FCD (n= 17, mean= 30.4, SD= 14.7, range= 12-50) compared to the normative sample (n= 401, mean= 48.8, SD= 11.2, range= 0-80) (t=6.54, df=416, p<0.0001). MMQ-Satisfaction scores amongst FCD participants (n=17, mean= 26.9, SD= 11.9, range= 12-56) were also significantly lower, indicating greater dissatisfaction with and concern about memory performance than the normative sample (n=401, mean= 43.9, SD=13.7, Range= 7-72) (t=5.04, df=416, p<0.0001).

### Predictors of Local and Global Metacognition

For the 17 patients who completed questionnaire measures, two regressions were performed using the hierarchical model for memory M-ratio: one with metacognitive bias as the sole independent variable, and one with 5 covariates taken from the questionnaire measures (GAD-7, PHQ-9 and each MMQ subscale). The HDIs for our estimations of regression betas for each predictor all contained zero and therefore did not provide evidence for linear relationships (**Table 3**). However, given the small numbers of trials and subjects entering into this regression, we note a relatively strong underlying relationship would have been needed to override the hierarchical group prior on zero effect.

**Table 3.** Hierarchical multiple regression predicting memory metacognitive efficiency

Predictors of Local and Global Metacognition
|  |  |  |
| --- | --- | --- |
| **Attribute** | **Mean sampled beta** | **HDI<sup>a</sup>** |
| *Regression 1 (5 covariates)* |  |  |
| GAD-7 | 0.018 | [-0.040, 0.074] |
| PHQ-9 | 0.011 | [-0.049, 0.074] |
| MMQ-Satisfaction | -0.008 | [-0.036, 0.020] |
| MMQ-Strategies | 0.003 | [-0.020, 0.024] |
| MMQ-Ability | 0.010 | [-0.011, 0.030] |
| *Regression 2 (1 covariate)* |  |  |
| Metacognitive bias | -0.00014 | [-0.0057, 0.0057] |
| Abbreviations: PHQ-9= Patient Health Questionnaire-9; GAD-7= Generalised Anxiety Disorder-7; MMQ= Multifactorial Memory Questionnaire. |  |  |
| <sup>a</sup>Sampled beta 95% Highest Density Interval. A 95% HDI which did not span zero would provide evidence that the effect of that predictor on metacognitive efficiency differed significantly from zero. |  |  |

Simple linear regressions were used to evaluate predictors of global metacognition (**Fig. 3**). More severe depression as measured by PHQ-9 was significantly associated with lower global SPEs as defined by the MMQ-Ability scores (β= −1.40 (SE=2.56), q= 0.01). Greater satisfaction about memory performance was significantly associated with higher global SPEs (β= 0.75 (SE=0.25), q= 0.02) while a greater use of strategies was significantly associated with lower global SPEs (β= −0.91 (SE=0.21), q= 0.01) (**Table 4**).

**Table 4.**
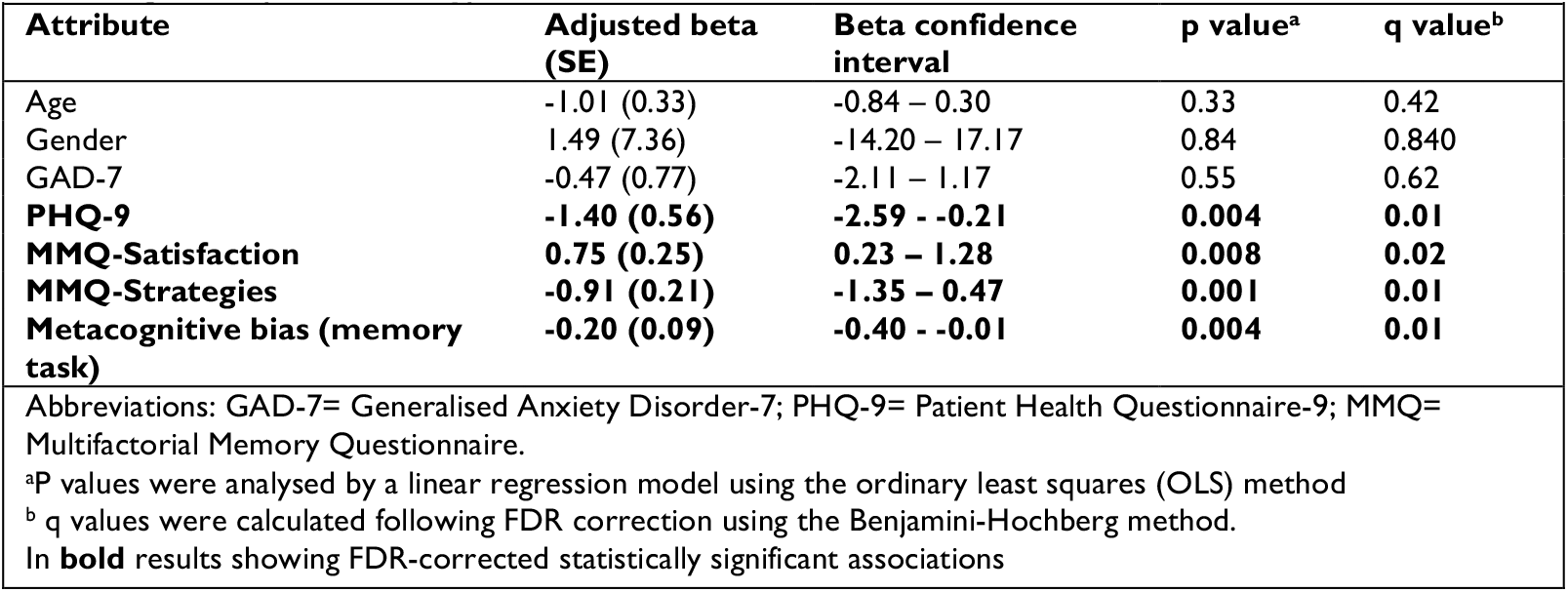
Association of demographic factors, mood and metacognitive measures with global metacognition (MMQ-Ability)

|  |  |  |  |  |
| --- | --- | --- | --- | --- |
| **Attribute** | **Adjusted beta (SE)** | **Beta confidence interval** | **p value<sup>a</sup>** | **q value<sup>b</sup>** |
| Age | -1.01 (0.33) | -0.84 – 0.30 | 0.33 | 0.42 |
| Gender | 1.49 (7.36) | -14.20 – 17.17 | 0.84 | 0.840 |
| GAD-7 | -0.47 (0.77) | -2.11 – 1.17 | 0.55 | 0.62 |
| **PHQ-9** | **-1.40 (0.56)** | **-2.59 – -0.21** | **0.004** | **0.01** |
| **MMQ-Satisfaction** | **0.75 (0.25)** | **0.23 – 1.28** | **0.008** | **0.02** |
| **MMQ-Strategies** | **-0.91 (0.21)** | **-1.35 – -0.47** | **0.001** | **0.01** |
| **Metacognitive bias (memory task)** | **-0.20 (0.09)** | **-0.40 – -0.01** | **0.004** | **0.01** |
| Abbreviations: GAD-7= Generalised Anxiety Disorder-7; PHQ-9= Patient Health Questionnaire-9; MMQ= Multifactorial Memory Questionnaire. |  |  |  |  |
| <sup>a</sup>P values were analysed by a linear regression model using the ordinary least squares (OLS) method |  |  |  |  |
| <sup>b</sup> q values were calculated following FDR correction using the Benjamini-Hochberg method. |  |  |  |  |
| In **bold** results showing FDR-corrected statistically significant associations |  |  |  |  |

**Figure 3.**
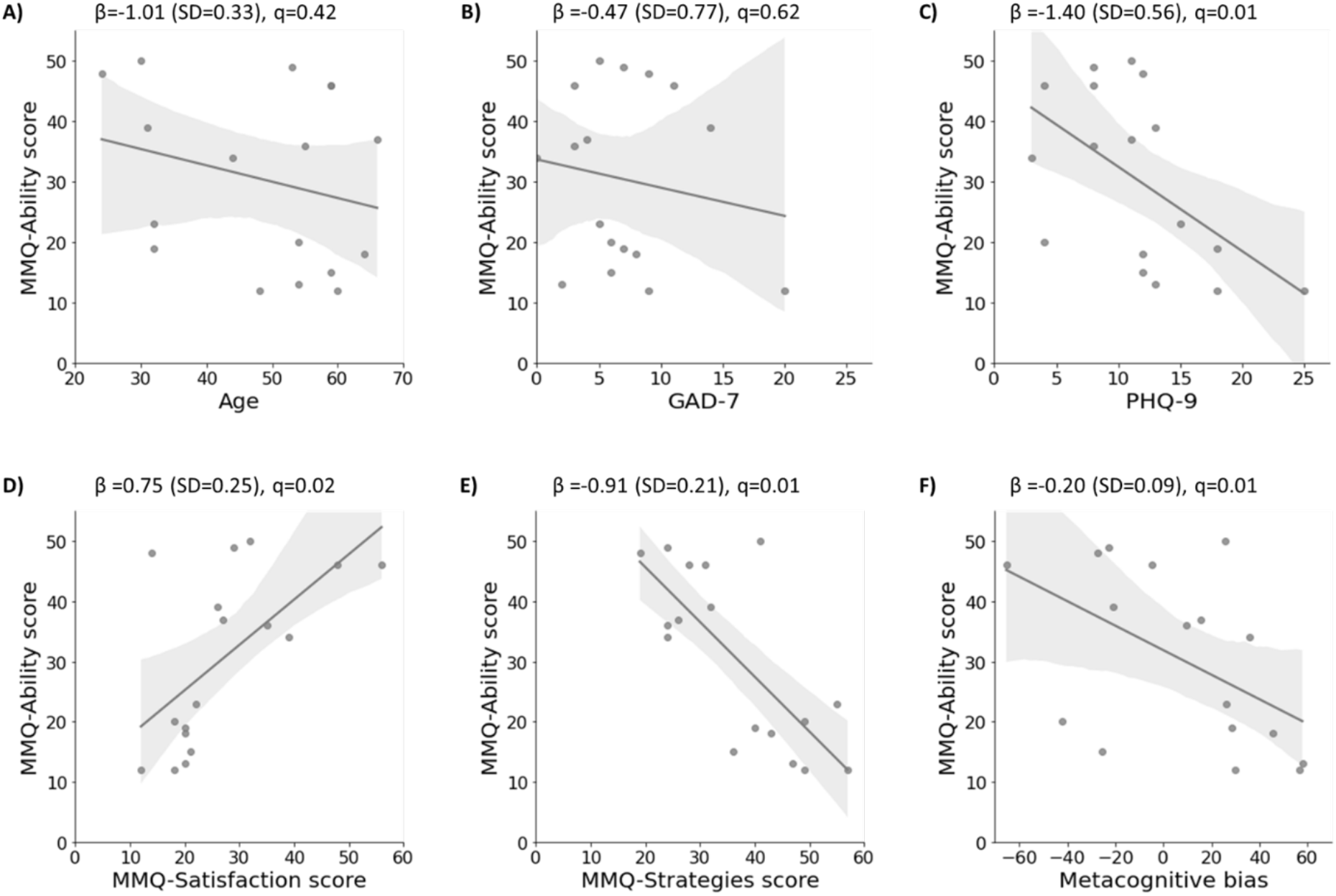
Relationship of age, affective symptomatology and local metacognitive measures with global metacognition (MMQ-Ability score). Regression plots for FCD participants with MMQ-Ability score as the dependent variable (y-axis) and the following independent variables; A) Age; B) GAD-7; C) PHQ-9; D) MMQ-Satisfaction score; E) MMQ-Strategies score; F) Metacognitive bias in the memory task. β coefficient values are presented along with q values that were calculated following FDR correction using the Benjamini-Hochberg method. Abbreviations: GAD-7= Generalised Anxiety Disorder-7; PHQ-9= Patient Health Questionnaire-9; MMQ= Multifactorial Memory Questionnaire. n=17 as one participant did not complete the MMQ questionnaire.

### Relationship between Local and Global metacognition

Local metacognitive bias in the memory task was significantly associated with MMQ-Ability (β= −0.20 (SE=0.09), q= 0.01) suggesting that higher local confidence estimates in the memory task were unexpectedly associated with lower global SPEs about memory.

## Discussion

Dysregulated metacognitive processing has been postulated as a unifying mechanism in FCD.^1,5-8^ A key finding in FCD is self-reports of low confidence in cognitive abilities in the absence of objective deficits, meaning that people with FCD explicitly report a metacognitive symptom. This study therefore sought to characterise local and global metacognitive processing in a sample of FCD patients. We found evidence for a dissociation between local and global metacognition: in FCD participants, local metacognitive efficiency was intact, while global metacognition was impaired. Additionally, in FCD, more strongly positive local metacognitive bias and higher depression scores were significantly associated with poorer global metacognition.

In our study, local metacognitive performance in FCD was similar to metacognitive efficiency in samples of healthy subjects.^19,20^ Metacognitive efficiency for memory tasks was higher than that in visual perception tasks as has been reported previously,^20,30^ which may be due to control processes playing a greater role on decision confidence in memory compared to visual perception tasks and/or differences in the structure of the underlying evidence (perceptual, mnemonic) entering the decision process.^30^

FCD patients had significantly lower self-appraisals of memory ability (lower self-performance estimates) as well as less satisfaction about their memory compared to existing normative data. This is consistent with previous work,^7,11^ which also used self-reported questionnaires to measure global metacognition.

Our study found intact local but impaired global metacognitive processing in patients with FCD. This disconnect between local and global metacognitive processing went against our primary hypothesis that both of these levels of metacognition would be impaired in FCD. However, it is possible that this disconnect is the very essence of cognitive internal inconsistency which is a canonical feature of FCD.^2^ An alternative explanation for our findings is that a person who is overly concerned about their cognitive performance might routinely attend more closely to and monitor that performance, so that they would ultimately develop effective local metacognitive discrimination that is indistinguishable from controls.

A disconnection between local and global metacognition can be explained using a Bayesian model of functional neurological disorder.^31^ Tailoring the model to FCD, this model posits an abnormal “prior” that contributes to the decoupling of top-down predictions of cognitive performance and bottom-up sensory information about actual cognitive performance. Local metacognition describes the ability to dynamically monitor moment-to-moment cognitive performance, thereby playing a crucial role in evaluating performance on individual decisions.^32^ This would correspond to the bottom-up sensory component within such a Bayesian model.

Meanwhile, global metacognition is characterised by the construction of priors or expectations at higher hierarchical levels and if impaired, this prior may lead to predictions of poor cognitive performance. In other words, an abnormal prior (global metacognition) may override bottom-up sensory input (local computations of confidence in individual decisions), giving rise to the perceived experience of impaired cognitive performance. In such a computational model,^31^ as has been postulated previously,^6^ if higher-order brain functions responsible for local metacognition are uncoupled from the predictions of the prior (shaped by global metacognition), the resulting cognitive difficulties may be perceived as being outside the subject’s control.

The relationship between local and global metacognition remains unclear, even in healthy subjects. We found that, in FCD participants, greater local confidence (metacognitive bias) was significantly associated with lower global metacognitive estimates. This was unexpected, and is in contrast to work in healthy participants which shows that confidence for individual decisions at a local metacognitive level is positively coupled to global SPEs in tasks that require learning about performance over time.^9^ In FCD, it is possible that this usage of local confidence signals to update global SPEs becomes disrupted, reflecting a pathological decoupling and loss of interaction between local and global metacognitive processing. Recent work using brain imaging suggests that a midline network spanning the ventromedial prefrontal cortex and precuneus may simultaneously represent both local and global confidence signals – indicating a potential anatomical locus for an interaction between these levels.^33^ Such a hypothesis could be further tested in FCD by employing tasks that examine learning of global SPEs over time.^9^ Global confidence estimates can also be incorporated as additional hierarchical levels within Bayesian models of metacognition. Mechanistically, such work could therefore also shed light on the mismatch between incorrect top-down predictions and bottom-up sensory input in the Bayesian model of FCD described above.

Another potential therapeutic approach would be to target improving global metacognition directly in FCD.^6^ Metternich and colleagues’ randomised controlled study^34^ demonstrated that a group psychological intervention comprising cognitive restructuring and psychoeducation led to significant improvements in Memory Self-Efficacy (MSE) (derived from three subscales of the Metamemory in Adulthood Questionnaire (MIA),^35^ a measure of global metacognition. Interestingly, global metacognition was significantly improved when measured at six months follow up but not immediately at the end of the intervention, suggesting that it may take time for cognitive restructuring to take place. This delay may conceal the effectiveness of the intervention from clinicians and could contribute to early termination because of lack of perceived efficacy of some psychological treatments.

Self-reported symptoms of depression but not anxiety were significantly associated with impaired global metacognition within the FCD group. The increased negative cognitions^21^ and neuroticism^36^ that people with depression experience may extend to their appraisal of their own cognitive performance. It might be expected that as an individual’s level of depression increases, their self-appraisal of cognitive performance will become more pessimistic and inaccurate, thereby leading to a reinforcing cycle. Clearly, not everyone with depression presents with global metacognitive deficits but when they arise co-morbidly there is likely to be a synergistic effect.^37^

### Limitations

The two key variables of interest, local and global metacognition were compared to existing data sets of healthy controls, which were not matched for gender or psychiatric co-morbidities and only one of the control groups was matched for age. In terms of age, participants in one of the comparison groups for local metacognition were significantly younger than our study participants,^20^ although there is some evidence that local metacognitive efficiency may worsen with age.^38^ MMQ-Ability, which we used to measure global metacognition, has a negligible relationship with age and gender,^26^ and so not having a gender and age matched comparison group for global metacognition is unlikely to have influenced our results.

Normative samples were also not matched for levels of depression or anxiety symptoms. This may be relevant in the analysis of local metacognition, because previous work has demonstrated that symptom dimensions related to depression and anxiety are associated with slightly higher metacognitive efficiency scores.^39^

The measurement of global metacognition relied on the use of the MMQ which is a self-reported questionnaire raising the possibility of response bias. In this regard, the development of behavioural experiments and analysis pipelines that enable global metacognition to be quantified objectively would be timely.^9^

Finally, FCD is an aetiologically heterogenous disorder.^1,37,40,41^ The small sample size in this study meant that it was not possibly to classify participants with different subtypes of FCD,^1,41^ and it remains possible that there may be variability in metacognition between these subgroups. Additionally, there is a risk of sampling bias given that participants were recruited from tertiary neuropsychiatric services and may not be representative of the significantly wider population of FCD patients who are managed in primary and secondary care.^1^

### Conclusions

Our findings suggest a decoupling of metacognitive processes in people with FCD wherein local metacognition is intact but global metacognition is impaired. Future work should incorporate recently developed behavioural experiments and analysis pipelines that evaluate how local metacognition influences global metacognition.^9^ A potential mechanistically plausible treatment may focus on re-establishing links between local and global metacognition.

In the meantime, given the lack of consensus on treatment in clinical practice, a psychological intervention focussed on improving global metacognition through cognitive restructuring and psychoeducation may be beneficial for FCD patients.

## Acknowledgements

We thank Dr Jorge Morales for providing full and swift access to data re-analysed here, as well as to relevant analysis code.

## Funding

RB is supported by a Wolfson-Eisai Clinical Research Training Fellowship. RH is supported by the NIHR UCLH BRC. AM is supported by the Mental Health and Justice Project funded by the Wellcome Trust (203376/2/16/Z). The Wellcome Centre for Human Neuroimaging is supported by core funding from the Wellcome Trust (203147/Z/ 16/Z). SMF is supported by a Sir Henry Dale Fellowship jointly funded by the Wellcome Trust and the Royal Society (206648/Z/17/Z). JH is funded by a Wellcome Clinical Research Career Development Fellowship (214547/Z/18/Z). For the purpose of Open Access, the author has applied a CC-BY public copyright license to any author accepted manuscript version arising from this submission.

## Competing Interests

The authors report no competing interests.

## Notes

### Competing Interest Statement

The authors have declared no competing interest.

